# Contrastive Self-supervised Learning for Neurodegenerative Disorder Classification

**DOI:** 10.1101/2024.07.03.24309882

**Authors:** Vadym Gryshchuk, Devesh Singh, Stefan Teipel, Martin Dyrba, ADNI, AIBL, FTLDNI study groups

## Abstract

Neurodegenerative diseases such as Alzheimer’s disease (AD) or frontotemporal lobar degeneration (FTLD) involve specific loss of brain volume, detectable in vivo using T1-weighted MRI scans. Supervised machine learning approaches classifying neurodegenerative diseases require diagnostic-labels for each sample. However, it can be difficult to obtain expert labels for a large amount of data. Self-supervised learning (SSL) offers an alternative for training machine learning models without data-labels. We investigated if the SSL models can applied to distinguish between different neurodegenerative disorders in an interpretable manner. Our method comprises a feature extractor and a downstream classification head. A deep convolutional neural network trained in a contrastive self-supervised way serves as the feature extractor, learning latent representation, while the classifier head is a single-layer perceptron. We used N=2694 T1-weighted MRI scans from four data cohorts: two ADNI datasets, AIBL and FTLDNI, including cognitively normal controls (CN), cases with prodromal and clinical AD, as well as FTLD cases differentiated into its sub-types. Our results showed that the feature extractor trained in a self-supervised way provides generalizable and robust representations for the downstream classification. For AD vs. CN, our model achieves 82% balanced accuracy on the test subset and 80% on an independent holdout dataset. Similarly, the Behavioral variant of frontotemporal dementia (BV) vs. CN model attains an 88% balanced accuracy on the test subset. The average feature attribution heatmaps obtained by the Integrated Gradient method highlighted hallmark regions, i.e., temporal gray matter atrophy for AD, and insular atrophy for BV. In conclusion, our models perform comparably to state-of-the-art supervised deep learning approaches. This suggests that the SSL methodology can successfully make use of unannotated neuroimaging datasets as training data while remaining robust and interpretable.

## 1. Introduction

Neurodegenerative diseases such as Alzheimer’s disease (AD) and frontotemporal dementia (FTD) are characterized by specific brain volume loss, which can be assessed in-vivo using structural magnetic resonance imaging (MRI). The usual radiological evaluation of MRI scans is mainly performed via visual examination, which is often time-consuming. Assistance systems for the automated detection of disease-specific patterns could be useful for better clinical diagnosis, as they can significantly decrease the evaluation time for the radiologists and neurologists, and help them focus on relevant brain regions. Convolutional neural networks (CNNs) models can automatically identify neurodegenerative diseases from MRI scans and achieve state-of-the-art results in medical imaging tasks. Recent development in the CNN architectures have in turn shaped the neuroimaging community, which is interested in automatic discovery of image features pertinent to neurological illnesses. Various tasks, such as - disease diagnosis, pathology localisation, anatomical region segmentation etc., now rely on the use of CNNs Dyrba et al. (2021); Qiu et al. (2020); Eitel et al. (2021); Wen et al. (2020); Han et al. (2022). CNN models are primarily trained in a *supervised* manner by using an external ground-truth label. Generating such labels for data samples is often burdensome and costly. Furthermore, CNN models require a large amount of training data to achieve competitive results. Such large datasets are not easily available within the medical domain due to the high cost of data collection and rarity of experts for annotations.

These constraints led us to reconsider the training of CNN models in a *supervised* manner, and to explore *self-supervised learning (SSL)* approaches. The SSL methods learn without any sample labels by utilizing the internal structure of the data, generating representative features. Architectures trained in a self-supervised manner are biologically plausible, provide extensive feature space, and compete with supervised approaches Orhan et al. (2020). We employed contrastive learning, a type of SSL methods, that allows learning of generalizable features from data by contrasting similar and different data samples.

The main goal of our study was to develop a computational approach for learning salient features from structural MRI data, enabling better generalization and interpretability. We hypothesized that SSL methods could learn meaningful structural representations, and resulting models could have comparable performances to supervised models. Therefore, in this paper, we propose to train a CNN model on structural MRI data within an SSL setup and then to evaluate this trained CNN model on a downstream classification task. We also explore a saliency mapping technique for highlighting relevant input regions. The main research question was defined as: *How does contrastive SSL paradigm compare against supervised learning paradigm in terms of predictive power? Are the models trained in contrastive self-supervised way on neuroimaging data interpretable?*

Our main contributions are: i) The conceptualization of a contrastive SSL architecture for the use with neuroimaging data, ii) A comparison between models trained on structural MRI data, using self-supervised and supervised approaches, in terms of their classification power, showing the results of our self-supervised model comparable to the supervised methods.

## 2. Background

### 2.1. Self-supervised learning

In recent years, we have seen an emergence of self-supervised learning (SSL) methods, which learn generalizable features without any data labels or ground truth information. By solving such an initial auxiliary task, they are then used for specific downstream tasks, e.g., identification of neurodegenerative disorders.

Models trained under the SSL approach have found application in different domains, i.e., image processing Jing and Tian (2020), video processing Schiappa et al. (2023), audio processing Liu et al. (2022a). Within the imaging domain, multiple auxiliary or so-called ‘pre-text’ tasks have been suggested previously: identifying data augmentations Reed et al. (2021); Chen et al. (2020), rotation prediction Chen et al. (2019), patch position prediction Doersch et al. (2015); Noroozi and Favaro (2016); Wei et al. (2019), image colorization Larsson et al. (2017, 2016), and contrastive learning Jaiswal et al. (2020).

SSL methods could be thought of as an alternative to pretraining or automated feature learning step and are related to the way how young children learn Orhan et al. (2020). Particularly, contrastive SSL methods try to learn the general structure present within the data, by using *supervisory signals* extracted from the data itself independently of the ground truth for any specific use-case.

#### 2.1.1. Formal definition of contrastive SSL

Contrastive learning tasks have received considerable attention within the SSL methods. Contrastive learning tasks aim at learning a latent space where embeddings of similar data samples are pulled together, and the embeddings of dissimilar data samples are pushed apart Gutmann and Hyvärinen (2010); Weng (2021); Chopra et al. (2005). Various loss functions have been suggested in order to increase the quality of learned embeddings, and expedite the training. These include: contrastive loss Gutmann and Hyvärinen (2010), triplet loss Chechik et al. (2010); Schroff et al. (2015), N-pair loss Sohn (2016), InfoNCE loss Oord et al. (2019) and Neighbourhood-based loss Sabokrou et al. (2019) etc. Contrastive learning is based on the use of positive and negative data pairs Grill et al. (2020); Chen et al. (2020), where a *positive pair* (*i, j*) consists of two similar data instances or views. In many studies, a data sample is paired with its own augmented variations in order to create such positive pairs. A *negative pair* generally contains two different data samples. The contrastive loss *𝓁* for a positive pair is formally defined as follows:

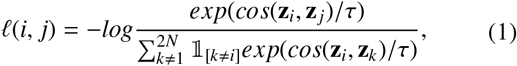

where τ is a scaling factor called temperature, 𝟙 is an indicator function with output values being 0 or 1, *N* is the number of training samples, exp(·) is the exponential function, and cos(·) is the cosine similarity function.

The Nearest-Neighbor Contrastive Learning (NNCLR) method Dwibedi et al. (2021) extends the common contrastive loss by keeping a record of recent embeddings of augmented views in a queue *Q*. Thus, the pairs are not directly compared, rather a projection embedding that is most similar to a view is selected from *Q* for the comparison with another view. The NNCLR contrastive loss *𝓁*_*n*_ is defined as:

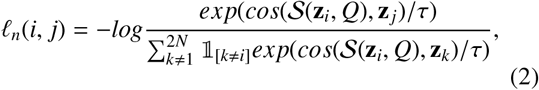

where *S* (**z**, *Q*) is the nearest neighbour function:

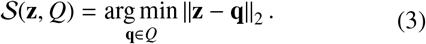

#### 2.1.2. Self-supervised learning in medical imaging

Compared to supervised approaches, there are relatively few applications of SSL methods in the medical domain Shurrab and Duwairi (2022); Huang et al. (2023). Most SSL approaches applied to MRI data focus on image reconstruction or segmentation Taleb et al. (2020); Hu et al. (2021a). Here, we briefly review two SSL methods applied by Taleb et al. (2020) within the medical imaging domain similar to our study. First, the *3D Contrastive Predictive Coding* operates on 3D data by splitting it into patches. It uses the InfoNCE loss for training the model, on an auxiliary task of predicting the latent representations of the next patches. Second, the *3D Exemplar networks* uses an auxiliary task of data augmentation prediction, where the model optimises the triplet loss.

Another methodology for SSL from MRI data was proposed by Hu et al. (2021a) that is based on parallel training of two networks with the objective of minimizing the reconstruction loss. Other recent research applied SSL to longitudinal AD MRI datasets in order to i) study methods for combining information from multiple imaging modalities Fedorov et al. (2021), or ii) to predict the trajectories of cognitive performance and/or cognitive decline Ouyang et al. (2021); Zhao et al. (2021).

### 2.2. Convolutional neural network backbones

Convolutional neural networks (CNN) have been the state-of-the-art solutions for computer vision tasks for almost a decade. In the last few years, numerous approaches on the advancement of CNNs were proposed: introduction of skip connections He et al. (2016); Huang et al. (2017), experimentation with model hyperparameters such as kernel size Ganjdanesh et al. (2023), normalisation strategies Ioffe and Szegedy (2015) and activation functions Dubey et al. (2022); Apicella et al. (2021), depthwise convolutions Howard et al. (2017), and model’s block architecture Sandler et al. (2018).

With the introduction of attention priors, the vision transformers (ViT) Dosovitskiy et al. (2020) soon became a viable alternative to the purely convolutional models, and currently represent the state-of-the-art model architecture as generic vision backbones. ViTs were inspired from the transformer models applied for the language processing tasks. To the best of our knowledge, there weren’t attempts of systematically comparing attention priors with convolutional priors. However, in their study Liu et al. (2022b) culminated many of the CNN advancements proposed over the years, and compared the resulting ConvNeXt model with comparable vision transformers. ConvNeXt Liu et al. (2022b) was proposed as a purely convolutional model, which achieved favourable results on common vision benchmarks such as the ImageNet Deng et al. (2009) and the COCO Lin et al. (2014) datasets, sometimes even providing higher accuracy than competing ViT models. Notably, ConvNeXt achieved these results while maintaining the computational simplicity and efficiency of standard CNN models, highlighting the importance of convolutional priors for vision tasks.

### 2.3. Feature attribution

With the growing popularity of CNN models and these models becoming the off-the-shelf baselines, there has also been a growing need to understand them. Multiple studies have attempted to explain and interpret the black-box CNN models. Within the domain of explainable AI (XAI), there are various methods to derive the importance of input features, i.e., the importance scores with respect to each prediction. These importance scores can be visualized by superimposing them on the input scans Van der Velden et al. (2022). Certain favoured methods of importance scoring are Layer-wise Relevance Propagation (LRP) Montavon et al. (2019); Kohlbrenner et al. (2020), Gradient-weighted Class Activation Mapping (Grad-CAM) Selvaraju et al. (2020), and Integrated Gradients (IG) Sundararajan et al. (2017). There have been multiple studies mapping importance scores to input regions, particularly within the neuroscience application of dementia detection Dyrba et al. (2021); Singh and Dyrba (2023); Böhle et al. (2019).

## 3. Methods

### 3.1. Neuroimaging datasets

We used T1-weighted brain MRI scans from publicly available neuroimaging repositories. The data scans in our study were pooled from the following data repositories: i) the Alzheimer’s Disease Neuroimaging Initiative (ADNI)^1^, study phases ADNI2 and ADNI3, ii) the Australian Imaging, Biomarker & Lifestyle Flagship Study of Ageing (AIBL)^2^ Ellis et al. (2009), collected by the AIBL study group, and iii) the Frontotemporal Lobar Degeneration Neuroimaging Initiative (FTLDNI)^3^.

In our study, the cognitively normal (CN) scan samples were consolidated from all three data cohorts. The samples with dementia due to Alzheimer’s disease (AD) and mild cognitive impairment (MCI) were collected from ADNI and AIBL data cohorts. While, FTLDNI was the only data cohort with samples categorised into different frontotemporal lobar degeneration (FTLD) sub-types, i.e., the behavioral variant of frontotemporal dementia (BV), the semantic variant of frontotemporal dementia (SV), and the progressive non-fluent aphasia (PNFA).

We applied the ‘t1-linear pipeline’ of the Clinica library Routier et al. (2021); Wen et al. (2020) to preprocess the raw MRI scans. The pipeline uses the N4ITK method for bias field correction and the SyN algorithm from ANTs to perform an affine registration for the alignment of each scan to the Montreal Neurological Institute (MNI) reference space. During the execution of the pipeline, some MRI samples were excluded due to quality checking or some missing information. Additionally, each scan was cropped to the size of 169 × 208 × 179 voxels with 1 mm isotropic resolution.

After applying preprocessing methods, our study includes 841 scans from the ADNI2, 968 scans from the ADNI3, 612 scans from AIBL and 273 scans from FTLDNI. Table 1 summarizes the sample statistics of the different data sources.

**Table 1:** Sample statistics of study data per diagnosis state. CN: a cognitively normal state, AD: dementia due to Alzheimer’s disease, MCI: mild cognitive impairment, BV: behavioral variant of frontotemporal dementia, SV: semantic variant of frontotemporal dementia, PNFA: progressive non-fluent aphasia, *μ*: mean, *σ:* standard deviation, MMSE: mini-mental state examination, F: female, M: male.

|  | CN | AD | MCI |  |
| --- | --- | --- | --- | --- |
| ADNI3 |  |  |  |  |
| Age: $\mu(\sigma)$ | 74 (7) | 77 (8.3) | 74.6 (8) | |
| MMSE: $\mu(\sigma)$ | 29.4 (0.7) | 20.8 (4.5) | 27.9 (1.1) | |
| Sex: F/M | 312/221 | 52/70 | 140/173 |  |
| ADNI2 |  |  |  |  |
| Age: $\mu(\sigma)$ | 75.8 (7) | 76.2(7.6) | 74.6 (7.9) | |
| MMSE: $\mu(\sigma)$ | 29.3 (0.7) | 21.1(4.3) | 27.8 (1.1) | |
| Sex: F/M | 110/94 | 120/163 | 151/203 |  |
| AIBL |  |  |  |  |
| Age: $\mu(\sigma)$ | 73.5 (6.4) | 75.4 (7.9) | 76.6 (6.5) | |
| MMSE: $\mu(\sigma)$ | 29.2 (0.8) | 19.5 (5.8) | 27.2 (1.3) | |
| Sex: F/M | 239/182 | 51/37 | 41/62 |  |
|  | CN | BV | SV | PNFA |
| FTLDNI |  |  |  |  |
| Age: $\mu(\sigma)$ | 64.3 (7.1) | 62.1 (5.8) | 62.7 (6.8) | 68.9 (7.7) |
| MMSE: $\mu(\sigma)$ | 29.7 (0.5) | 22.6 (6.2) | 22.5 (5.7) | 24.9 (5.5) |
| Sex: F/M | 72/58 | 23/48 | 14/23 | 19/16 |

### 3.2. Proposed self-supervised learning pipeline

Our proposed method consists of two modules: a feature extractor and a classification head. The feature extractor is a convolutional neural network trained without any sample labels in a self-supervised manner. The classification head is a simple neural network subsequently trained in a supervised way. The proposed architecture is shown in Figure 1.

**Figure 1:**
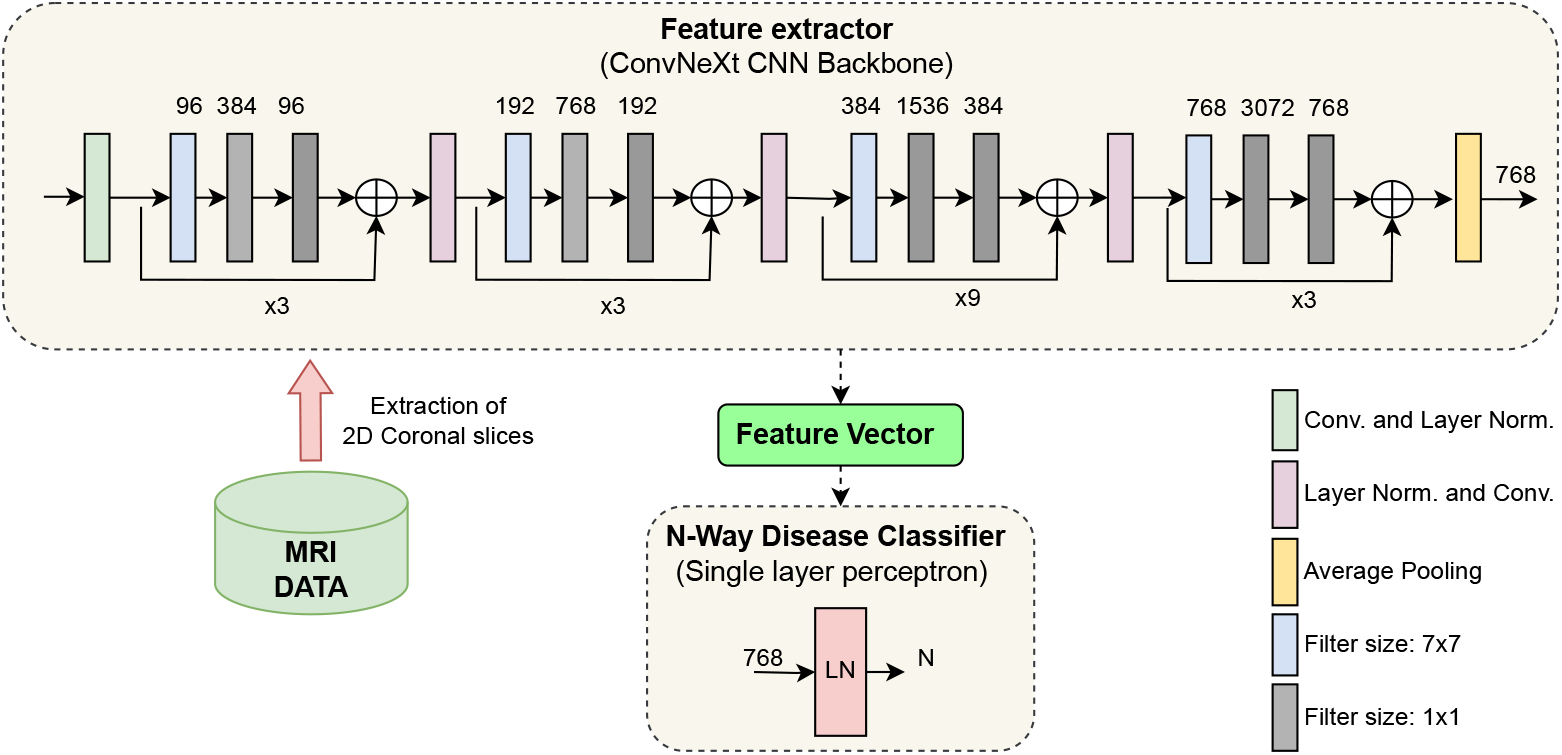
Illustration of the proposed architecture. (Top) ConvNeXt, a CNN model, trained under a self-supervised learning paradigm, extracts features from coronal brain slices. (Bottom center) The classification head learns to classify neurodegenerative disorders from the extracted features. CNN: convolutional neural network. LN: layer normalization. Conv: convolutional operation. LN: Layer Normalization

After executing the t1-linear pipeline of the Clinica library, we obtained a 3D image for the brain of each participant. However, we only used 2D convolutional operations, as they reduce the CNN parameter space and model complexity. We selected only the coronal plane for the present study. In each MRI sample, there were in total 208 coronal slices, however, we considered only 120 coronal slices in the middle. The slices from the middle contain the relevant regions, such as the hippocampus and the temporal lobe, which are reported to be affected already in the earliest stages of Alzheimer’s disease Whitwell et al. (2008).

#### Feature extractor

We used the ConvNeXt model Liu et al. (2022b) as the backbone for the SSL framework. It was trained with the NNCLR loss *𝓁*_*n*_ for learning visual representations from the input data (see Eq. 2). We applied a series of random augmentations to a randomly selected coronal slice for the creation of positive pairs, as exemplified in Figure 2.

**Figure 2:**
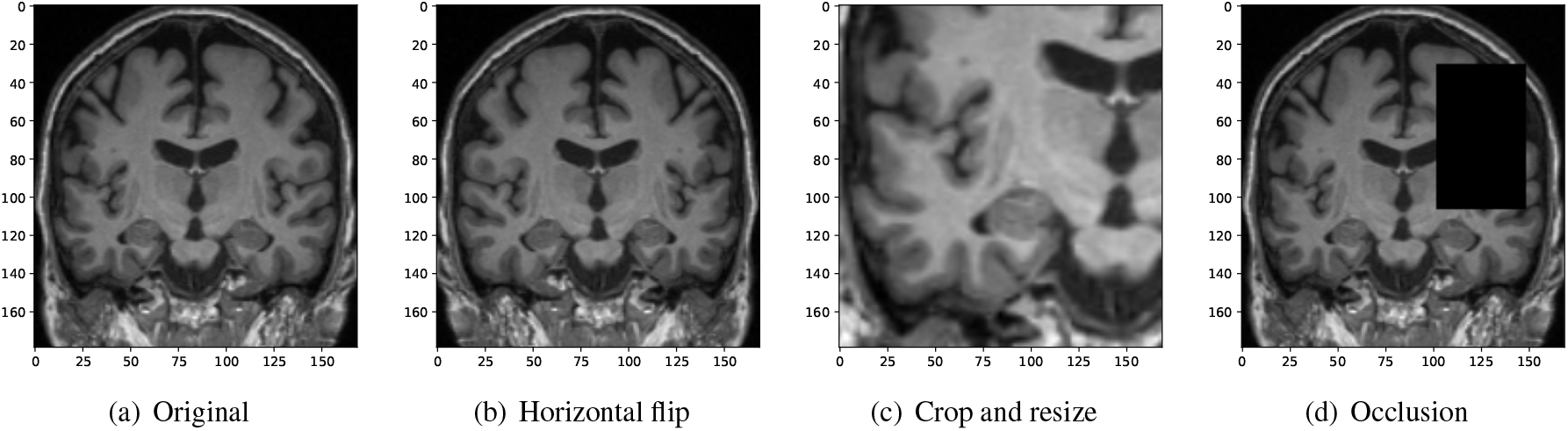
Randomly applied data augmentations to the input during training.

The loss optimised for a data batch was:

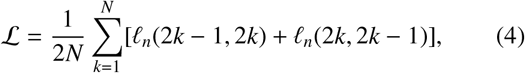

where *𝓁*_*n*_ is the NNCLR loss from Eq. 2, 2*k* − 1 and 2*k* represent the indices of the same augmented slice, and *N* is the total number of training samples.

Specifically, we used the ‘tiny’ ConvNeXt model variant Liu et al. (2022b) as our backbone model. It has a configuration with sequential blocks set to (3, 3, 9, 3) and the number of output channels equalling to (96, 192, 384, 768). ConvNeXt culminates many architectural advancements such as larger kernel sizes of 7×7, skip connections, inverted bottleneck, Gaussian error linear units (GELU) as activation function, layerwise normalization (LN) strategy instead of batch normalisations (BN), etc. The ConvNeXt model and pretrained model weights can be downloaded from the publicly available PyTorch library^4^.

#### Classification head

While using the ConvNeXt model as a feature extractor, we considered the output produced by a 2*D* adaptive average pooling layer after the last convolutional block as input for the subsequent ‘classification head’ (Figure 1). That means the classification head takes as input the latent feature representations of the MRI scans that where processed by the backbone CNN model. The dimension of the extracted feature vector per MRI slice is 768. Our classification head is a simple neural network consisting of a single fully-connected layer preceded by a layer normalization operation (Figure 1).

### 3.3. Feature attribution

Integrated gradients (IG) can be applied to various data modalities, such as texts, images or structured data Sundararajan et al. (2017). IG was chosen over other feature attribution methods because of its strong theoretical justifications, such as the completeness property of the integrated gradients. IG considers a straight path from some baseline to the input, and computes the gradients along that path. These accumulated gradients are called integrated gradients. However, this accumulation is an approximation of the actual integration of the gradients, and the number of steps taken between the baseline to the input determines the quality of this approximation. To calculate the IG importance scores, a mean CN image was used as the baseline to the IG attribution method. We used the IG implementation provided by the Captum library^5^ to calculate importance maps for the MRI scans with respect to the classification task.

### 3.4. Experimental Setup

#### Training the feature extractor

We trained a feature extraction model (ConvNeXt) using the NNCLR method on the ADNI3, ADNI2, and FTLDNI data for three learning trials. For each trial, we created random training and test sets. These sets were held constant for all further experiments. If more than one MRI recording was available per participant, then we assigned all of the participant’s MRI scans only to one set, thus avoiding data leakage. This resulted in 10% of data belonging to the test set.

The model was trained for 1000 epochs using a batch size of 180 samples. The size of the NNCLR queue *Q* was set to 8192. We applied three different data augmentation techniques with a probability of 0.5 to produce views visualized in Figure 2(b-d): horizontal flip, cropping and resizing, and occlusion. We experimented with different data sources for training the feature extractor, i.e., utilising in-domain medical images vs. training with out-of-domain natural images. More details about model training and results could be found in the supplementary.

#### Training the classification head

To determine if a 3D MRI scan belongs to a specific diagnostic group, we first derive the latent representation vectors for coronal 2D slices using the ConvNeXt feature extractor, and then make a prediction for each slice using the classification head. For evaluation with the test data, we applied a simple voting procedure, in which the most frequently occurring group label determined the final group assignment. We trained the classification head for 100 epochs, on the same three training trials that were used to train the feature extractors. We use a batch size of 64 samples and decayed the learning rate with cosine annealing after every 20 epochs.

We experimented with various setups for training a classification head, with a) keeping the weights of the feature extractor frozen vs. unfrozen, i.e., letting the weights change during the classification head training, and b) for the downstream task we compared different multi-class classification heads, i.e., predicting four (CN, MCI, AD, BV) or three classes - (CN, MCI, AD) and (CN, AD, BV), and binary classification heads - (CN, AD), (CN, BV), and (AD, BV). Furthermore, we evaluated our models on the independent AIBL dataset, which was not used during training. The independent test dataset enabled us to assess the generalizability of our approach.

## 4. Results

### 4.1. Diagnostic group separation

We evaluated the manner in which the classification head could be setup. We compared multi-class vs. binary classification heads. Table 2 shows the achieved results of our proposed architecture for the identification of neurodegenerative disorders, using a frozen ConvNeXt feature extractor trained under NNCLR SSL pradigm on the brain images. The numbers reported were averaged over threee learning trials. For the binary (AD vs. CN) classification model, the balance accuracy reached 82% for the cross-validation test sets and 80% for the independent AIBL data cohort. In Section 5.1 below, we discuss the achieved results and compare them with the state of the art.

**Table 2:** Classification results of our proposed architecture, consisting of a frozen feature extractor trained under a SSL paradigm, and a single-layer neural network as the downstream classification head. In a multi-class setup, micro averages are reported for the sensitivity and specificity metrics. CN: a cognitively normal, AD: dementia due to Alzheimer’s disease, MCI: mild cognitive impairment, BV: behavioral variant of frontotemporal dementia, MCC: Matthews correlation coefficient.

|  | Balanced accuracy | MCC | Sensitivity | Specificity |
| --- | --- | --- | --- | --- |
| **Cross-validation test set (ADNI2/3 and FTLDNI)** |  |  |  |  |
| AD vs. MCI vs. CN vs. BV: | 0.60±0.03 | 0.32±0.02 | 0.51±0.01 | 0.84±0.00 |
| AD vs. MCI vs. CN: | 0.56±0.02 | 0.32±0.03 | 0.55±0.02 | 0.78±0.01 |
| AD vs. CN vs. BV: | 0.78±0.03 | 0.55±0.05 | 0.73±0.02 | 0.87±0.01 |
| BV vs. CN: | 0.88±0.03 | 0.57±0.03 | 0.90±0.08 | 0.86±0.02 |
| AD vs. CN: | 0.82±0.04 | 0.61±0.08 | 0.82±0.05 | 0.82±0.03 |
| AD vs. BV: | 0.93±0.01 | 0.73±0.04 | 0.85±0.02 | 1.00±0.00 |
| **Independent test set (AIBL)** |  |  |  |  |
| AD vs. MCI vs. CN: | 0.53±0.01 | 0.30±0.03 | 0.69±0.01 | 0.84±0.01 |
| AD vs. CN: | 0.80±0.01 | 0.59±0.01 | 0.66±0.02 | 0.94±0.01 |

### 4.2. Model interpretability

To highlight the input regions that were found to be useful by the SSL model, we used the Integrated Gradients (IG) attribution method. IG calculates the importance scores for the input regions for a specified prediction label. The IG importance scores were calculated for every sample of the test data set (from ADNI2/3 and FTLDNI), on which our multi-class model (AD vs. CN vs. BV) makes a correct classification. Figure 3 presents mean IG importance scores for the disease types AD and BV, visualised over the brain scan of a healthy sample chosen from the ADNI cohort. While making a prediction towards the diseased classes, the red regions in the image highlight input regions representing the evidence for the diseased class, while the green regions in the image highlight input regions representing the evidence against the diseased class. The mean importance scores were thresholded to visualize the most relevant findings.

**Figure 3:**
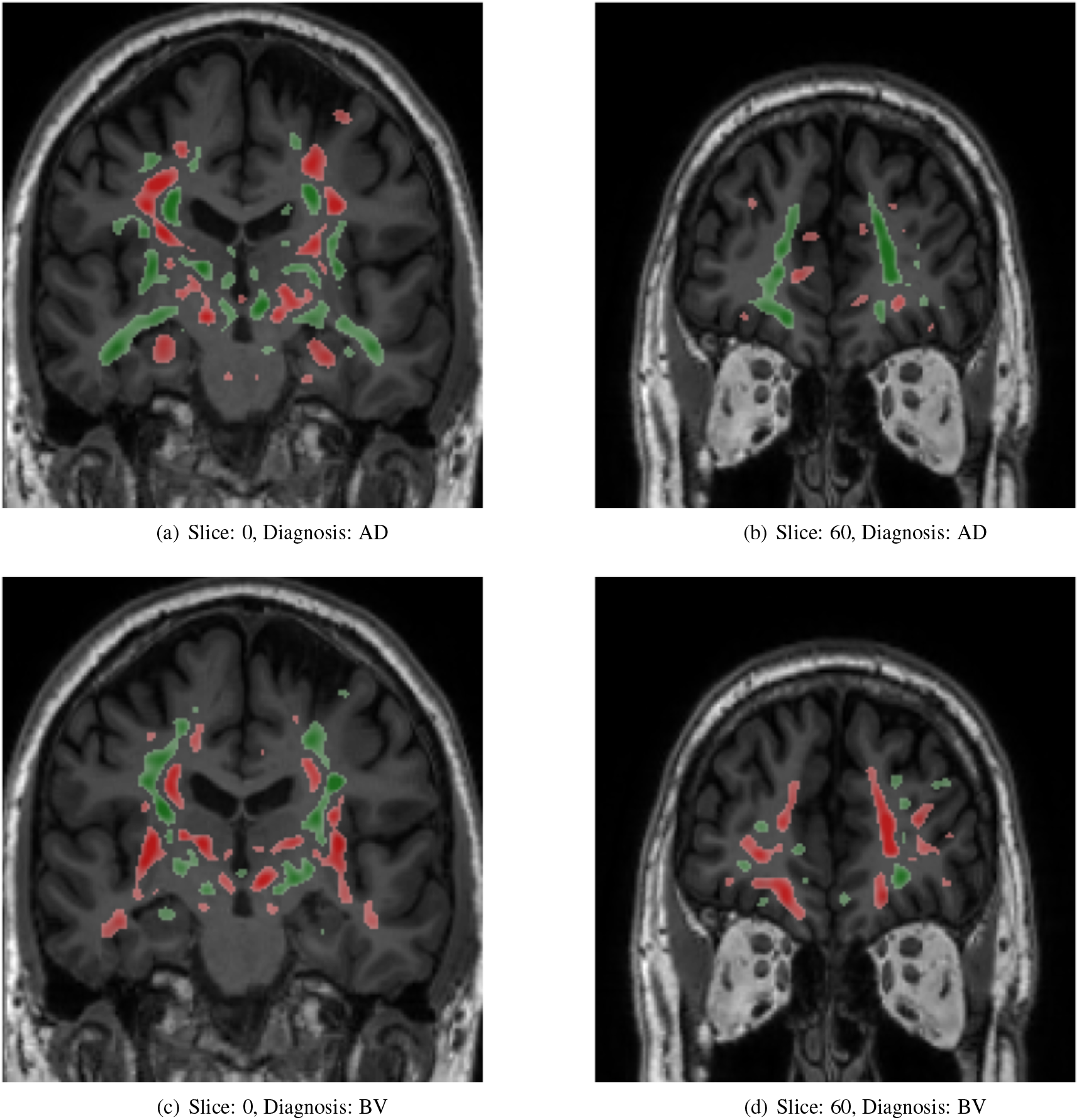
Mean attribution maps derived from the Integrated Gradients method for correctly identified AD and BV samples. Green and red color highlight pixel contributions to the model’s prediction. Here, red highlight evidence for the respective disease classification and green indicates evidence against it. The attribution map overlay image was smoothed and thresholded to highlight relevant findings and improve visualization. AD: dementia due to Alzheimer’s disease, BV: behavioral variant of frontotemporal dementia.

## 5. Discussion

### 5.1. Feature learning

In our proposed SSL framework, we rely on signals that are derived from the data itself rather than on external classification target labels to train a feature extractor. We trained our SSL model while restricting input to a subset of 2D coronal slices. It should be noted that other SSL studies also avoided training 3D CNN with high input resolution, and followed similar 2D approaches as our study Couronné et al. (2021) or alternatively needed to downscale the 3D images to a very low 64 × 64 × 64 resolution Ouyang et al. (2021); Fedorov et al. (2021).

In Table 2, upon comparing results from various settings of classification heads trained over a frozen feature extractor, we can observe a general trend, i.e., the binary classification for separating cognitively normal (CN) and Alzheimer’s disease (AD) samples is a much simpler task than the multi-class classification of CN, mild cognitive impairment (MCI), AD and behavioral variant of frontotemporal dementia (BV) samples. This finding has often been reported in other studies in the field.

In the multi-class classification setting, the AD vs. MCI vs. CN model, often confuse MCI samples with CN or AD samples. This reflects the progressive nature of the Alzheimer’s dementia, with MCI being intermediate stage between CN and AD. Interestingly, we found that the AD vs. MCI vs. CN vs. BV model is substantially better at separating BV samples from the other CN, MCI and AD samples, with the recall (=sensitivity) of the BV class being 0.89, compared to the average micro recall of the same model being 0.51. This finding points towards different underlying pathologies of different dementia diseases - frontotemporal dementia and AD. The same fact could also be corroborated from the high performance metrics of the binary AD vs. BV model.

Our AD vs. CN vs. BV multi-class model achieves a balanced accuracy of 78%. Certain supervised methods solve the same task, achieving performance metrics as - Ma et al. (2020) reports (simple) accuracy of 85.97% from a model comparable to ours and 88.28% from a model with more data information sources and generative data augmentation, and Hu et al. (2021b) reports (simple) accuracy of 66.79% on a larger diverse dataset, and 91.83% on a smaller cleaner dataset. While our BV vs. CN binary model achieves a balanced accuracy of 88.2%. For the same task Moguilner et al. (2023) reports (simple) accuracy of 80% and 95% on MRI scans with 1.5T and 3T strength, respectively.

The closest work to ours are by Ouyang et al. (2021) that reported group separation results using a SSL approach. However, they explicitly models longitudinal aspect of the data for the disease trajectory prediction task, these design choices explicitly encodes more information in the model than our models. On the ADNI dataset, they achieved a balanced accuracy of 81.9% from a multilayer perceptron classifier with frozen feature encoder and 83.6% post fine-tuning the feature encoder, for the binary AD vs. CN task. While, similarly our model with a frozen feature extractor, achieves a balanced accuracy of 82% on ADNI dataset. For the same downstream binary classification task - AD vs. CN, on an independent test set (AIBL), our model achieves a balanced accuracy of 80%, which is only a two percentage point drop from the cross-validation testing of the model, highlighting the robustness of the model.

In Table 3, we compare our model evaluation results with the state-of-the-art studies that also used AIBL as an independent test dataset. Here, we compare our SSL model with other models trained in a supervised manner. Qiu et al. (2020), reports manual expert rating scores, with a simple accuracy metric of 82.3%. This performance level is comparable to that of our SSL models, which achieved the simple accuracy measure of 89.9% on the AIBL independent test set. It should be noted that some papers did not report the *balanced accuracy* measure, thus, their ‘simple’ accuracy results might be biased towards the majority class of cognitively normal people who comprise 80% in the AIBL dataset for the group comparison AD vs. CN.

**Table 3:** Comparison of our proposed method with the state-of-the-art. The results are provided for studies that used the AIBL dataset for independent evaluation and the group comparison AD vs. CN. In some studies balanced accuracy was not reported, ‘simple’ accuracy is provided instead, which might be biased towards the majority class (=CN). AD: dementia due to Alzheimer’s disease, CN: cognitively normal. SSL: Self-supervised learning, SL: Supervised learning. CNN: Convolutional neural network.

| Study training<br>on the ADNI dataset | Method details | Balanced accuracy |
| --- | --- | --- |
| Our method | SSL, 2D slice-level CNN | 0.797±0.009 |
| Wen et al. (2020) | SL, 2D slice-level CNN | 0.756±0.015 |
| Wen et al. (2020) | SL, 3D patch-level CNN | 0.802±0.016 |
| Wen et al. (2020) | SL, 3D subject-level CNN | 0.862±0.016 |
| Dyrba et al. (2021) | SL, 3D subject-level CNN | 0.832±0.030 |
|  |  | Simple accuracy |
| Our method | SSL, 2D slice-level CNN | 0.899±0.003 |
| Qiu et al. (2020) | SL, 3D patch-level CNN | 0.870±0.022 |
| Han et al. (2022) | SL, 3D subject-level CNN | 0.865 |
| Han et al. (2022) | SL, 3D patch-level CNN | 0.875 |
| Qiu et al. (2020) | Expert Neurologists | 0.823±0.094 |

With regard to our achieved level of performance, we can conclude that the ConvNeXt model trained under a SSL paradigm learns generalizable features for the subsequent downstream classification tasks without requiring data sampling techniques or sophisticated data augmentations, and consequently achieving competitive results in comparison to other supervised approaches. The reported results show that our model learnt meaningful feature representations, in a self-supervised manner, which helped it in successfully separating different dementia stages and types.

### 5.2. Neural network interpretability

We chose the SSL paradigm for extracting more generalizable image features independently of a downstream task. However, the SSL paradigm also allows the backbone model to learn features of the brain that may correlate with a specific neurodegenerative disorder.

We applied the Integrated Gradients (IG) method to interpret the models and provide insights into the significance of input regions for the predictions. The IG importance scores were calculated for samples from the test dataset for which our AD vs. CN vs. BV multiclass model makes correct classifications. Figure 3 illustrates the mean IG importance scores for classifying samples into the AD or BV group. In Figure 3(a-b), we see the hippocampus region being highlighted in red, for the AD classification. Temporal lobe atrophy, specifically hippocampus atrophy, is a hallmark sign of Alzheimer’s disease. In Figure 3(c-d), we see the insula and frontal lobe regions being highlighted in red. Insular atrophy is associated with behavioral variant frontotemporal dementia Moguilner et al. (2023); Seeley (2010); Luo et al. (2020); Mandelli et al. (2016). It is of great interest to see the IG maps separately highlighting regions, which in literature are often associated with AD and BV pathology.

Notably, the model successfully learned to not consider tissue outside of the brain or regions outside of the skull. However, the derived attributions provide a rather general indication of important input regions throughout the brain including primarily the grey and white matter tissue. Few studies have pointed out the complex nature of IG importance scores highlighting multiple image features, both for and against a class instance, which makes their comprehension non-trivial Adebayo et al. (2018); Kakogeorgiou and Karantzalos (2021).

### 5.3. Limitations and future work

Our study uses only a subset of coronal slices for making sample level classifications. We acknowledge that the selection of the full slice set along the coronal axis or selection of the full 3D MRI data could have a positive effect on classification performance, however, the main goal of the study was to investigate the application of SSL and to compare it with traditional supervised approaches, thus only a subset of slices along the coronal axis were chosen as input. In addition, training 3D CNN requires a considerably larger parameter space than 2D models. Learning a 3D CNN is currently a computationally intractable problem for self-supervised learning, as it relies on - a) very large data corpus, b) data augmentation algorithms which are markedly more computationally expensive in 3D, and c) many learning iterations as training typically converges much slower than in supervised learning. More specifically, training our models for 1000 epochs on a single NVIDIA Quadro RTX 6000 GPU took on average 1 day and 3 hours.

In future, for training better feature extractors, we would try to incorporate more spatial neuroanatomical information, by combining three CNNs, i.e., one trained along each orthogonal planes - axial, coronal, and sagittal, and hence learning a feature representations for full 3D MRI data, as was recently proposed for supervised models Qiao et al. (2021). Alternatively, a Transformer model could be designed to efficiently process smaller 3D patches of the brain, which would not require separate CNN models for each plane Qiu et al. (2020); Wen et al. (2020); Han et al. (2022).

With regards to neural network interpretability and feature attribution, a comprehensive analysis of the salient features and feature attribution methods lies outside the scope of our current work. Further experiments are required to holistically understand our SSL model and produced more informative importance maps. In our future work, we will explore other methods for feature attribution, and methods to summarize attributions per brain region to assess if specific disease patterns emerge.

We also intend to include more data in our future studies, for learning more robust models. Specifically, we intend to add FTLD data cohorts.

## 6. Conclusion

We presented an architecture for the identification of neurodegenerative diseases from MRI data, consisting of a feature extractor and a classification head. The feature extractor used the ConvNeXt architecture as a backbone, which was trained under a self-supervised learning paradigm with nearest-neighbor contrastive learning (NNCLR) loss on brain MRI scans. The feature extractor model was used for subsequent downstream tasks by training only an additional single-layer neural network component which performs the classification. From our experiments, we show that CNN models trained under SSL paradigm have comparable performance to state-of-the-art CNN models trained in a supervised manner. With this presented approach, we provide a practical application of self-supervised learning on MRI data, as well as also demonstrate the application of attribution mapping methods for such systems in order to improve interpretability of the model’s decision.

## Data Availability

The data used for this study is publicly available and can be obtained from the respective repositories: Alzheimer's Disease Neuroimaging Initiative (ADNI) http://adni.loni.usc.edu/data-samples/access-data, Australian Imaging Biomarkers and Lifestyle flagship study of ageing (AIBL) https://aibl.csiro.au, and Frontotemporal Lobar Degeneration Neuroimaging Initiative (FTLDNI) https://memory.ucsf.edu/research-trials/research/allftd.

https://aibl.csiro.au

https://memory.ucsf.edu/research-trials/research/allftd

## Abbreviations

AD: Alzheimer’s disease
ADNI: Alzheimer’s Disease Neuroimaging Initiative
AIBL: Australian Imaging, Biomarker & Lifestyle Flagship Study of Ageing
BN: Batch normalisation
BV: behavioral variant of frontotemporal dementia
CN: Cognitively normal participants
CNN: Convolutional neural network
ConvNeXT: A highly optimized CNN model architecture recently introduced by Liu et al. (2022b)
DZNE: Deutsches Zentrum für Neurodegenerative Erkrankungen (German Center for Neurodegenerative Diseases)
FTLD: Frontotemporal lobar degeneration
FTLDNI: Frontotemporal Lobar Degeneration Neuroimaging Initiative
GELU: Gaussian error linear units
Grad-CAM: Gradient-weighted class activation mapping
IG: Integrated gradients
InfoNCE: A form a contrastive loss metric, where NCE stands for Noise-Contrastive Estimation
LN: Layer-wise normalization
LRP: Layer-wise relevance propagation
MCC: Matthews correlation coefficient
MCI: Mild cognitive impairment
MNI: Montreal Neurological Institute
MRI: Magnetic resonance imaging
NNCLR: Nearest-Neighbor Contrastive Learning
PNFA: Progressive non-fluent aphasia
SL: Supervised learning
SSL: Self-supervised learning
SV: semantic variant of frontotemporal dementia
ViT: Vision Transformers
XAI: Explainable artificial intelligence

## Declarations

## Acknowledgements

Data collection and sharing for this project was funded by the Alzheimer’s Disease Neuroimaging Initiative (ADNI) (National Institutes of Health Grant U01 AG024904) and DOD ADNI (Department of Defense award number W81XWH-12-2-0012). ADNI is funded by the National Institute on Aging, the National Institute of Biomedical Imaging and Bioengineering, and through generous contributions from the following: AbbVie, Alzheimer’s Association; Alzheimer’s Drug Discovery Foundation; Araclon Biotech; BioClinica, Inc.; Biogen; Bristol-Myers Squibb Company; CereSpir, Inc.; Cogstate; Eisai Inc.; Elan Pharmaceuticals, Inc.; Eli Lilly and Company; EuroImmun; F. Hoffmann-La Roche Ltd and its affiliated company Genentech, Inc.; Fujirebio; GE Healthcare; IXICO Ltd.;Janssen Alzheimer Immunotherapy Research & Development, LLC.; Johnson & Johnson Pharmaceutical Research & Development LLC.; Lumosity; Lundbeck; Merck & Co., Inc.;Meso Scale Diagnostics, LLC.; NeuroRx Research; Neurotrack Technologies; Novartis Pharmaceuticals Corporation; Pfizer Inc.; Piramal Imaging; Servier; Takeda Pharmaceutical Company; and Transition Therapeutics. The Canadian Institutes of Health Research is providing funds to support ADNI clinical sites in Canada. Private sector contributions are facilitated by the Foundation for the National Institutes of Health (www.fnih.org). The grantee organization is the Northern California Institute for Research and Education, and the study is coordinated by the Alzheimer’s Therapeutic Research Institute at the University of Southern California. ADNI data are disseminated by the Laboratory for Neuro Imaging at the University of Southern California. A complete listing of ADNI investigators can be found at https://adni.loni.usc.edu/wp-content/uploads/how_to_apply/ADNI_Acknowledgement_List.pdf.

Data collection and sharing for this project was funded by the Frontotemporal Lobar Degeneration Neuroimaging Initiative (National Institutes of Health Grant R01 AG032306). The study is coordinated through the University of California, San Francisco, Memory and Aging Center. FTLDNI data are disseminated by the Laboratory for Neuro Imaging at the University of Southern California.

Data used in the preparation of this article was also obtained from the Australian Imaging Biomarkers and Lifestyle flagship study of ageing (AIBL) funded by the Commonwealth Scientific and Industrial Research Organisation (CSIRO). The AIBL researchers contributed data but did not participate in analysis or writing of this report. AIBL researchers are listed at www.aibl.csiro.au. AIBL study methodology has been reported previously Ellis et al. (2009).

## Ethical approval

Data collection of the respective neuroimaging initiatives was approved by the internal review boards of each of the participating study sites. See https://adni.loni.usc.edu and https://aibl.csiro.au for details. All initiatives met common ethical standards in the collection of the data such as the Declaration of Helsinki. Analysis of the data was approved by the internal review board of the Rostock University Medical Center, reference number A 2020-0182.

## Availability of Data and Material

Our source code for data processing, model training and evaluation, and creating attribution maps will be made publicly available at: https://github.com/VadymV/clinic-net

## Authors’ contributions

VG: conceptualization, methodology, data curation and processing, coding and software development, visualization, and writing - original draft. DS: conceptualization, coding and software development, visualization, writing – original draft. MD: conceptualization, methodology, visualization, and writing - original draft, writing - review and editing, supervision. ST: conceptualization, methodology, writing - review and editing, supervision, and clinical validation.

## Footnotes

1 ADNI: https://adni.loni.usc.edu/

2 AIBL: https://aibl.csiro.au/

3 FTLDNI: https://memory.ucsf.edu/research-trials/research/allftd

4 ConvNeXt via PyTorch: https://pytorch.org/vision/main/models/convnext.html

5 Captum: https://captum.ai/

